# Utility of Cerebrospinal Fluid Protein Levels as a Potential Predictive Biomarker of Disease Severity in HIV-Associated Cryptococcal Meningitis

**DOI:** 10.1101/2023.12.10.23299793

**Authors:** John Kasibante, Eesha Irfanullah, Abduljewad Wele, Elizabeth Okafor, Kenneth Ssebambulidde, Samuel Okurut, Enock Kagimu, Jane Gakuru, Morris K. Rutakingirwa, Timothy Mugabi, Edwin Nuwagira, Samuel Jjunju, Edward Mpoza, Lillian Tugume, Laura Nsangi, Abdu K Musibire, Conrad Muzoora, Joshua Rhein, David B. Meya, David R. Boulware, Mahsa Abassi

**Affiliations:** Infectious Diseases Institute, College of health sciences, Makerere University. P.O. Box 22418, Kampala, Uganda; Division of Infectious Diseases & International Medicine, Department of Medicine, University of Minnesota, Minneapolis, MN 55455, USA; Department of Medicine, Mbarara University of Science and Technology, P.O Box 1410, Mbarara, Uganda

**Keywords:** HIV, cryptococcal meningitis, cerebral spinal fluid, CSF protein, CSF cytokines, predictive biomarker

## Abstract

**Background:** Cerebrospinal fluid (CSF) protein levels exhibit high variability in HIV-associated cryptococcal meningitis from being normal to markedly elevated. However, the clinical implications of CSF protein levels in cryptococcal meningitis remain unclear.

**Methods:** We analysed data from 890 adults with HIV-associated cryptococcal meningitis randomized into two clinical trials in Uganda between 2015 and 2021. CSF protein was grouped into ≥100 mg/dL (n=249) and <100 mg/dL (n=641). We described baseline clinical variables and mortality by CSF protein levels.

**Results:** Approximately one-third of individuals had a baseline CSF protein ≥100 mg/dL. Those with CSF protein ≥100 mg/dL were more likely to present with Glasgow coma scale scores <15 (P<0.01), self-reported seizures at baseline (P=0.02), higher CD4 T-cells (p<0.001), and higher CSF white cells (p<0.001). Moreover, those with a baseline CSF protein ≥100 mg/dL also had a lower baseline CSF fungal burden (p<0.001) and a higher percentage of sterile CSF cultures at day 14 (p=0.02). Individuals with CSF protein ≥100 mg/dL demonstrated a more pronounced immune response consisting of upregulation of immune effector molecules pro-inflammatory cytokines, type-1 T-helper cell cytokines, type-3 chemokines, and immune-exhaustion marker (p<0.05). 18-week mortality risk in individuals with a CSF protein <100 mg/dL was 34% higher, (unadjusted Hazard Ratio 1.34; 95% CI, 1.05 to 1.70; p=0.02) than those with ≥100 mg/dL.

**Conclusion:** In cryptococcal meningitis, individuals with CSF protein ≥100 mg/dL more frequently presented with seizures, altered mental status, immune activation, and favourable fungal outcomes. Baseline CSF protein levels may serve as a surrogate marker of immune activation and prognosis.

## Background

Sub-Saharan Africa is home to an estimated 20.1 million of the 38.4 million people living with HIV (PLWH) globally [1]. Since 2010, increased access to HIV testing, antiretroviral therapy (ART), and improved management of opportunistic infections has decreased HIV-associated mortality by 47% [1]. Despite advancements in HIV care and management, HIV-associated opportunistic infections remain a significant burden in sub-Saharan Africa, where 54% of cryptococcal meningitis and 63% of all cryptococcal-related mortality occur [2]. Cryptococcal meningitis mortality, even in the setting of clinical trials, remains unacceptably high, with 10-week mortality exceeding 25% [3, 4]. Therefore, there is a need to identify and understand the role of predictive biomarkers of cryptococcal meningitis severity to inform targeted interventions and improve outcomes.

Cerebrospinal fluid (CSF) proteins are characterized by marked intra-individual and inter-individual variability in healthy individuals with approximately 80% of protein originating from blood and 20% from the central nervous system [5]. Elevated CSF protein therefore indicates either a dysfunction in the blood-CSF or the blood-brain barrier [6]. In healthy individuals, CSF protein levels range between 15 and 60 mg/dL. In cryptococcal meningitis, CSF protein levels are highly variable, ranging from normal levels to as high as >20 times the upper limit of normal [6, 7]. In 1974, Diamond and Bennet found that in cryptococcal meningitis, baseline CSF protein levels were higher in persons who survived compared to those who died [8]. In two recent small retrospective studies, individuals with cryptococcal meningitis and high baseline CSF protein had a better response to antifungal therapy and improvement in clinical symptoms compared to those with lower baseline CSF protein levels [7, 9]. While existing data offers intriguing insights into the relationship between baseline CSF protein levels and outcomes in cryptococcal meningitis, it is crucial to acknowledge that the current evidence remains limited and inconclusive.

Our study aims to enhance our understanding of the role of CSF protein in cryptococcal meningitis and to assesses its potential as a predictive biomarker of severity in HIV-associated cryptococcal meningitis. Additionally, we sought to investigate the relationship between CSF protein levels with established risk factors of severe disease, such as seizures, altered mental status, CD4 T-cell count, CSF fungal burden, CSF opening pressure, and CSF cytokine profile [10-12]. Furthermore, we examined the predictive value of elevated CSF protein as a predictor of mortality.

## Methods

### Study Design

We carried out a prospective cohort study of Ugandans adults diagnosed with HIV-associated cryptococcal meningitis as a secondary analysis of two randomized clinical trials: (i) the Adjunctive Sertraline for the Treatment of HIV-associated Cryptococcal Meningitis (ASTRO-cm) trial enrolled from March 9, 2015, to May 29, 2017; and (ii) the AMBIsome Therapy Induction Optimisation (AMBITION-cm) trial enrolled from January 2018 through February 2021 [3, 4]. A detailed description of the study design for each clinical trial is published elsewhere [4, 13]. In brief, the ASTRO-cm trial investigated the impact of adjunctive sertraline on cryptococcal meningitis treatment outcomes. The AMBITION-cm trial investigated the efficacy of single high-dose liposomal amphotericin B on fluconazole and flucytosine backbone in cryptococcal meningitis. Participants were recruited and enrolled at Mulago National Specialist Hospital, Kiruddu National Referral Hospital, and Mbarara Regional Referral Hospital in Uganda. All participants diagnosed with a first episode of HIV-associated cryptococcal meningitis by a positive finger stick and CSF cryptococcal antigen (CrAg) were included in this analysis [14, 15]. All study participants had a diagnostic LP performed at baseline. CSF samples were collected and sent to the laboratory and analysed for CSF cell counts, quantitative fungal cultures, and protein concentration.

Ethical approvals for both clinical trials were granted from the Mulago Hospital Research and Ethics Committee, the Uganda National Council of Science & Technology, and the University of Minnesota Institutional Review Board. All participants or their surrogate provided their written consent for study participation.

### Cytokine quantification

We assessed CSF cytokine and chemokine concentrations at baseline among a subset of the ASTRO-CM study participants. CSF samples were centrifuged at 400g for 4 minutes at 4^°^C, supernatant was collected and stored at −80^°^C. Frozen samples were transported to the University of Minnesota and cytokines/chemokines were measured using the Human Luminex Discovery Assay (R&D systems, Minneapolis, MN, USA). The 43 cytokine and chemokine assay included **mediators of inflammatory response**: interleukin (IL) −1a, IL-1b, IL-6, platelet derived growth factor (PDGF)-AA, PDGF-AB, and vascular endothelial growth factor (VEGF); **type-1 T helper cell (Th1) associated cytokines**: macrophage inflammatory protein (MIP)1a/CCL3, MIP1b/CCL4, RANTES/CCL5, tumor necrosis factor (TNF)-α, IL-12, IL-15, interferon (IFN)-γ, interferon-inducible protein (IP)-10/CXCL10, granulocyte-macrophage colony stimulating factor (GM-CSF), and granzyme B; **Th2 cytokines**: IL-4, IL-5, IL-13, IL-25, IL-33, and eotaxin; **Th3 cytokines**: MIP3a/CCL20, growth-related oncogene (GRO)-α/CXCL1, GRO-β/CXCL2, CXCL8/IL-8, IL-17A, and granulocyte colony stimulating factor (G-CSF); **modulators of immune response**: IL-10, IL-1 receptor antagonist (IL-1ra), Endothelial growth factor (EGF), and transforming growth factor (TGF)-α; **immune exhaustion**: programmed death ligand-1 (PD-L1), **Type-1 interferon-**α and interferon-β, and **non-classified chemokines**: MIP3b/CCL19, IL-3, CD40 ligand/TNFSF5, fractalkine/CX3CL1, Flt3 ligand,

### Statistical methods

We initially divided patients into tertiles: low CSF protein (0 to < 45 mg/dL), middle CSF protein (≥45 mg/dL to <100 mg/dL), and high CSF protein (≥100 mg/dL). We found no significant difference between the low and middle tertile groups in terms of survival, and therefore we combined them into an aggregate CSF protein group of <100 mg/dL. We summarized baseline demographic variables and laboratory parameters by CSF protein <100 mg/dL vs. CSF protein ≥100 mg/dL. We also evaluated the impact of ART status at enrollment on CSF protein levels and its contribution to clinical presentation and outcomes given the role that ART plays in unmasking immune reconstitution inflammatory syndrome (IRIS). Data are presented as median with interquartile range (IQR) or proportions with statistical testing via Kruskal-Wallis’s test for medians and Fisher’s exact tests for proportions. P values <0.05 were considered statistically significant. We adjusted cytokine P-values with the Benjamini-Hochberg procedure to control the false discovery rate [16]. We compared survival by Kaplan-Meier curves and Cox proportion hazard regression analysis using both the adjusted and unadjusted models. The adjusted survival model included differentially distributed co-variates between groups, including CSF opening pressure >250 mmH_2_O, CSF white cell count, seizures, and CSF cryptococcal quantitative colony forming units (CFUs). Quantitative CFUs were log_10_ transformed for normalization. We conducted all analyses with SAS version 9.4 (SAS Institute, Cary, NC).

## Results

### Demographics

We included 890 adults with HIV-associated cryptococcal meningitis in our analysis. 72% (641/890) had CSF protein levels <100 mg/dL at baseline. We found no statistically significant differences in age, gender, and weight at baseline between CSF protein groups “Table 1”. While there was no difference between the percentage of persons who were on ART (p=0.82) or the duration of time on ART (p=0.05) at baseline, we found a statistically significant higher CD4 T-cell count in persons with CSF protein ≥100 mg/dL (median CD4 = 28, IQR 10 to 65 cells/μL) as compared to persons with CSF protein <100 mg/dL (median CD4 = 15, IQR 6 to 41 cells/μL; p<0.001). Individuals with CSF protein ≥100 mg/dL presented with more self-reported seizures (p=0.02) and a Glasgow Coma Scale score <15 (p<0.01) compared to those with CSF protein <100 mg/dL.

**Table 1.** Clinical Parameters and Treatment Outcomes by Baseline CSF Protein Levels.

|  | N | CSF Protein<br><100 mg/dL<br>(N = 641) | N | CSF Protein<br>≥100mg/dL<br>(N = 249) | P-value<sup>1</sup> |
| --- | --- | --- | --- | --- | --- |
| Age (years) | 641 | 35 [30, 40] | 249 | 35 [30, 42] | 0.46 |
| Female | 641 | 264 (41) | 249 | 87 (35) | 0.09 |
| Weight (kg) | 561 | 50 [46, 60] | 224 | 53 [47, 60] | 0.26 |
| **Antiretroviral Therapy (ART) Status** |  |  |  |  |  |
| Currently on ART | 640 | 307 (48) | 249 | 117 (47) | 0.82 |
| Months on ART | 306 | 4 [1, 32] | 115 | 3 [1, 13] | 0.05 |
| ≤14 Days | 306 | 51 (17) | 115 | 26 (23) | 0.10 |
| 15-30 Days |  | 37 (12) |  | 11 (10) |  |
| 31-90 Days |  | 45 (15) |  | 25 (22) |  |
| >90 Days |  | 173 (57) |  | 53 (46) |  |
| **Clinical** |  |  |  |  |  |
| Seizures | 641 | 92 (14) | 249 | 52 (21) | **0.02** |
| Glasgow Coma Scale <15 | 640 | 231 (36) | 249 | 114 (46) | **<0.01** |
| CD4 <50 cells/mm<sup>3</sup> categorical | 614 | 485 (79) | 240 | 158 (66) | **<0.001** |
| CD4 cells/mm<sup>3</sup> | 614 | 15 [6, 41] | 240 | 28 [10, 65] | **<0.001** |
| **CSF Opening Pressure mmH<sub>2</sub>O** |  |  |  |  |  |
| Baseline | 590 | 259 [170, 390] | 234 | 255 [170, 350] | 0.22 |
| Day 3 | 149 | 300 [196, 420] | 54 | 290 [210, 400] | 0.77 |
| Day 7 | 292 | 180 [110, 280] | 119 | 180 [100, 268] | 0.34 |
| Day 14 | 223 | 210 [130, 295] | 98 | 157 [100, 240] | **<0.01** |
| **Quantitative CSF Culture, log<sub>10</sub> CFU/mL** |  |  |  |  |  |
| Baseline | 622 | 4.68 [3.2, 5.5] | 247 | 4.09 [2.3, 5.3] | **<0.001** |
| Day 3 | 156 | 4.45 [3.1, 5.2] | 60 | 3.48 [2.2, 4.9] | **0.04** |
| Day 7 | 313 | 1.91 [0.0, 3.6] | 131 | 0.00 [0.0, 2.8] | **<0.01** |
| Day 14 | 245 | 0.00 [0.0, 1.7] | 102 | 0.00 [0.0, 1.2] | 0.38 |
| **CSF White Cell cells/mm<sup>3</sup>** |  |  |  |  |  |
| CSF <5 white cells/mm<sup>3</sup> | 636 | 460 (72) | 246 | 77 (31) | **<0.001** |
| Baseline | 636 | <5 [<5, 10] | 246 | 53 [<5, 150] | **<0.001** |
| Day 3 | 159 | <5 [<5, 30] | 60 | 28 [<5, 138] | **<0.001** |
| Day 7 | 312 | <5 [<5, 45] | 130 | 35 [<5, 95] | **<0.001** |
| Day 14 | 248 | <5 [<5, 25] | 103 | 25 [<5, 65] | **<0.001** |
| **CSF Sterility status** |  |  |  |  |  |
| Baseline Sterile Culture | 622 | 47 (8) | 247 | 28 (11) | 0.08 |
| Day 14 Sterile Culture | 429 | 191 (45) | 149 | 83 (56) | **0.02** |
| Early Fungicidal Activity, log<sub>10</sub> CFU/mL/d | 474 | 0.333 (0.186, 0.480) | 191 | 0.326 (0.208, 0.482) | 0.43 |

Data are represented as Median [IQR] or N (%)
<sup>1</sup>Kruskal-Wallis test for medians; Fisher Exact tests for proportions
Day 3, 7, and 14 values are collected within -/+1 day within the visit window

### CSF characteristics

Baseline CSF white cells were significantly higher in individuals with CSF protein ≥100 mg/dL (median 53, IQR <5 to 150 white cells/mm^3^ CSF) compared to those with CSF protein <100 mg/dL (median <5, IQR <5 to 10 white cells/mm^3^ CSF; p <0.001). CSF white cell counts remained significantly higher through days 3, 7, and 14 among individuals with CSF protein ≥100 mg/dL as compared to CSF protein <100 mg/dL (p<.001). Baseline CSF quantitative cryptococcal cultures were significantly lower in persons with CSF protein ≥100 mg/dL (median 12 300, IQR 200 to 200 000 CFU/mL) compared to persons with CSF protein <100 mg/dL (median 48 000, IQR 1585 to 316 000 CFU/mL; p<0.001). At day 14, the proportion of persons with a sterile CSF culture was higher in those with CSF protein ≥100 mg/dL compared to those with CSF protein <100 mg/dL (CSF protein ≥100 mg/dL 56% (83/149) vs. CSF protein <100 mg/dL 45% (191/429); p=0.02). We found no differences in baseline CSF opening pressure between individuals with CSF protein ≥100 mg/dL compared to CSF protein <100 mg/dL (p=0.22).

CSF cytokine assays revealed that individuals with CSF protein ≥100 mg/dL had significantly higher levels of pro-inflammatory cytokines, interleukin(IL)-1β, IL-6, PDGF-AA and VEGF; type-1 T helper cell (Th1) response cytokines IL-12, IL-15, interferon-γ, TNF-α, and GM-CSF; effector molecules granzyme B and TRAIL; type-2 T helper cell (Th2) response cytokine IL-5 and Eotaxin; type 3/ Th17 related cytokines IL-17, G-CSF, CXCL1, CXCL2, CXCL8, and CCL20; and immune exhaustion marker PD-L1; as compared to those with CSF protein <100 mg/dL “Table 2”.

**Table 2:**
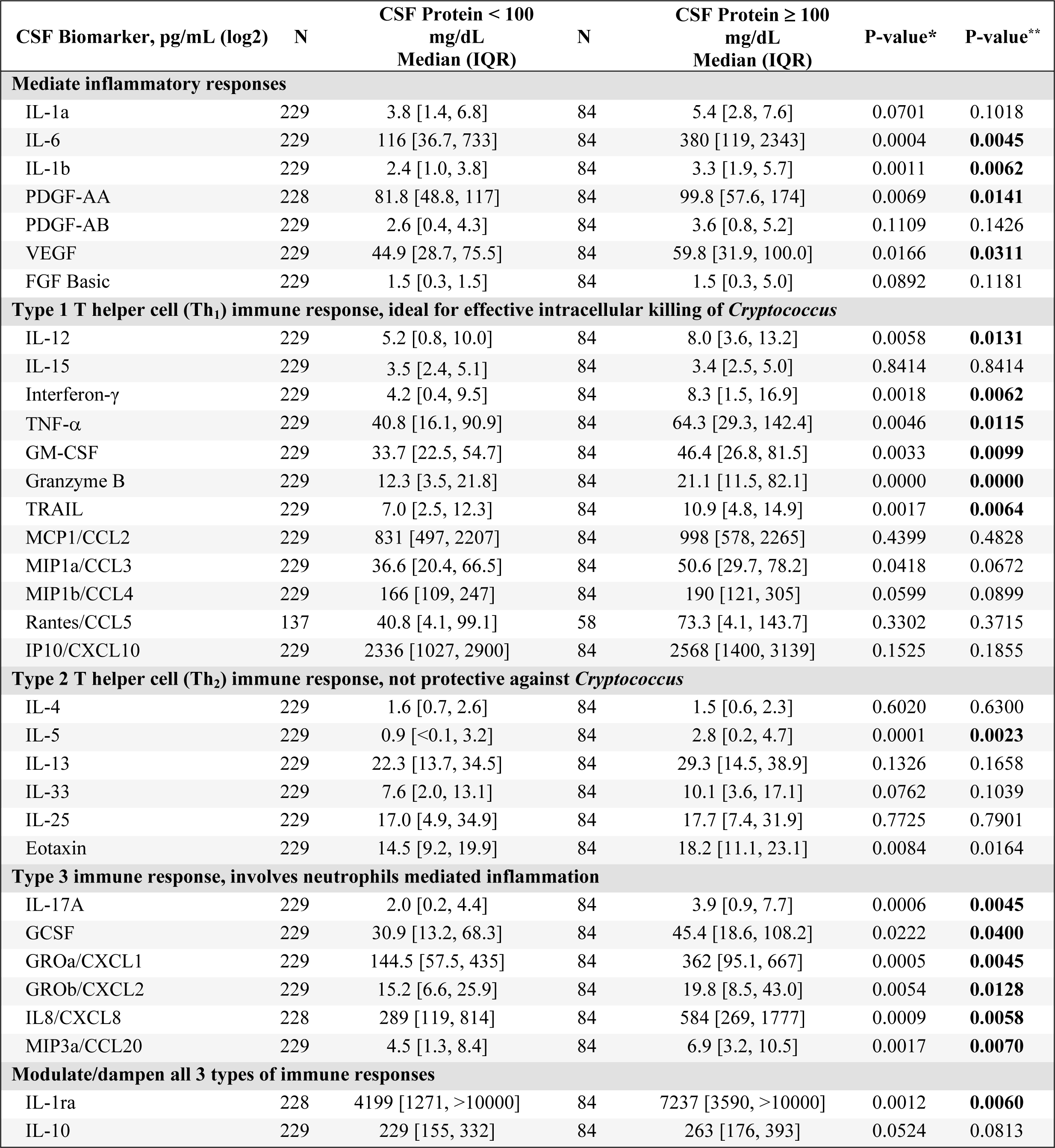
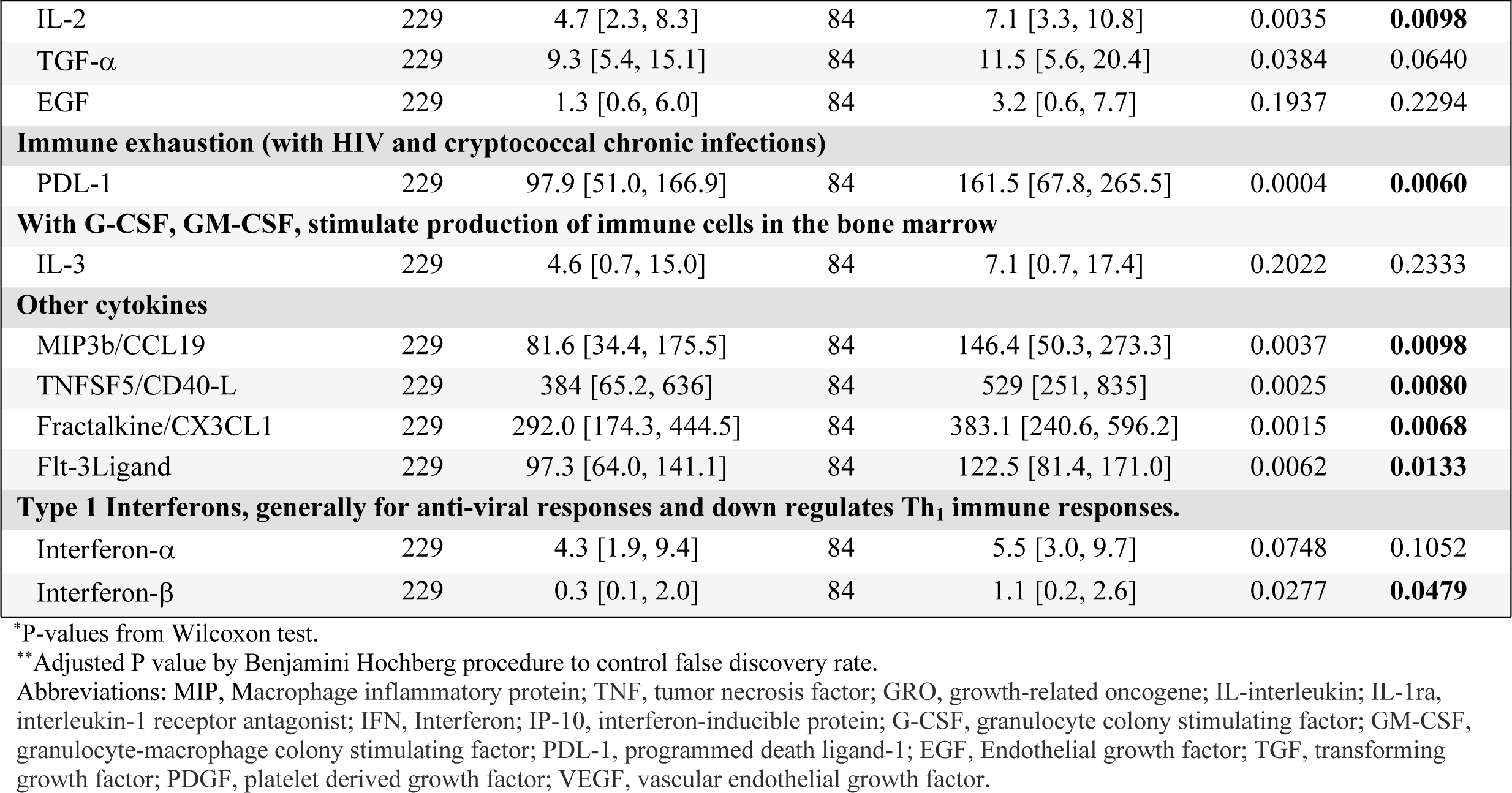
Baseline CSF Cytokines and Chemokines by Baseline CSF Protein Levels.

### Effect of ART on CSF Protein Levels

ART status at study enrolment influenced clinical parameters in both CSF protein categories “Table 3”. Individuals on ART and with a baseline CSF protein ≥100 mg/dL were more likely to present with a Glasgow coma scale score <15 as compared to persons on ART and with a baseline CSF protein <100 mg/dL (98/307 vs. 50/117, p<0.04). There was no difference in Glasgow coma scale score <15 between CSF protein groups in individuals who were not receiving ART on admission (p=0.10). Persons with a baseline CSF protein ≥100 mg/dL had a higher CD4 T-cell count, higher CSF white cell count at baseline and through day 14, and a lower baseline CSF fungal burden compared to persons with CSF protein <100 mg/dL, irrespective of baseline ART status. Individuals on ART with a CSF protein ≥100 mg/dL had a higher percentage of sterile cultures at day 14 compared to individuals on ART and with a CSF protein <100 mg/dL (p<0.01). There was no difference in the percentage of day 14 sterile cultures between CSF protein groups in those who were not on ART at baseline (p=0.78).

**Table 3:**
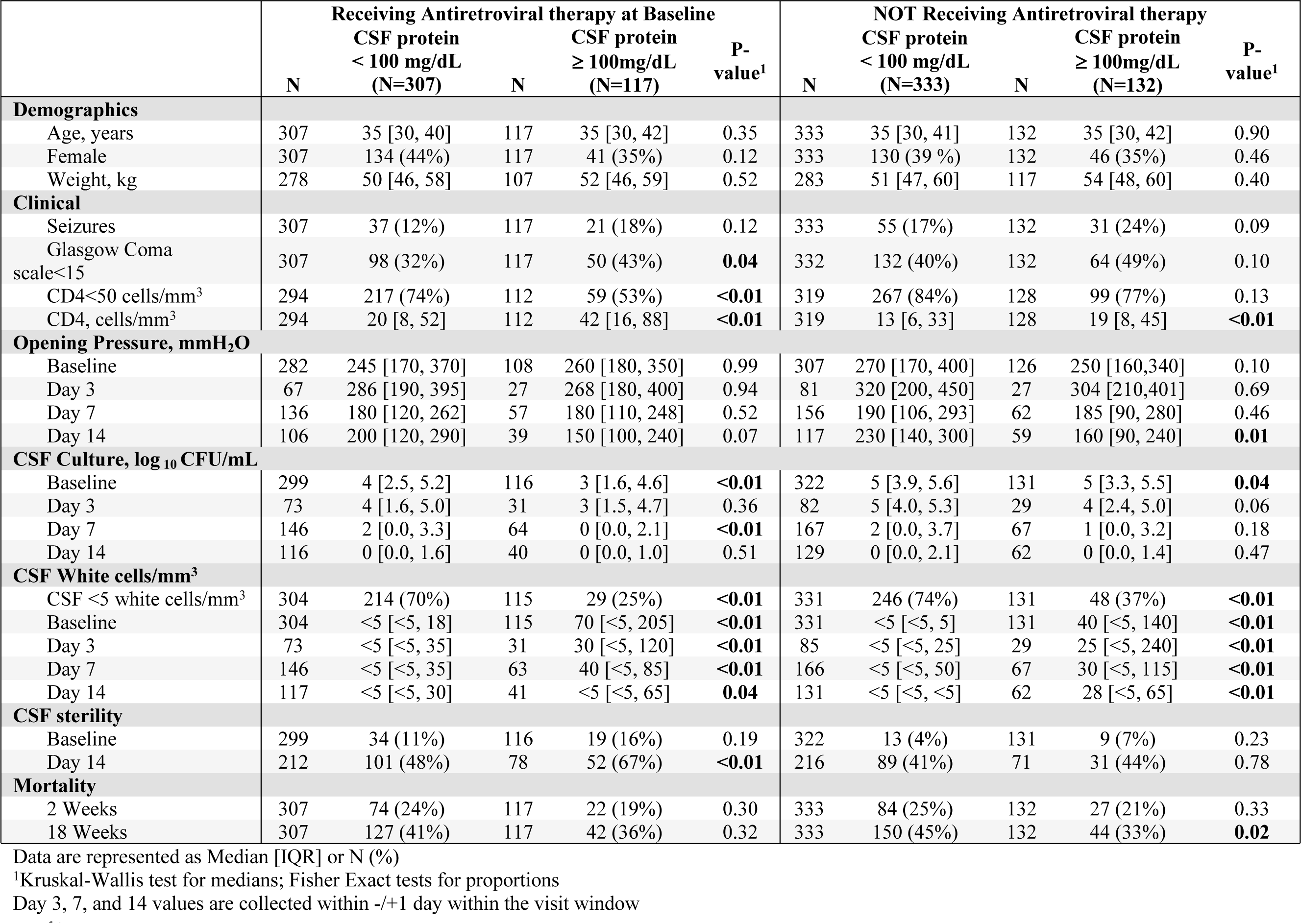
Clinical Presentation and Outcomes by Baseline CSF Protein Levels and ART Status.

### Outcomes

CSF protein levels influenced survival outcomes. Through 18 weeks, those with a CSF protein <100 mg/dL were at a 34% higher risk of death compared to individuals with CSF protein ≥100 mg/dl (Hazard Ratio=1.34; 95% CI 1.05 to 1.70; p=0.02) “Table 4, Fig 1a”. Adjustment for baseline CSF fungal burden, Glasgow coma scale score <15, CSF white count, CSF opening pressure, and baseline self-reported seizures, attenuated the 18-week mortality risk between CSF protein groups (adjusted Hazard Ratio = 1.24; 95%CI, 0.94 to 1.64; p=0.13). Mortality rates through 18-weeks differed by ART status. Mortality was highest among individuals who were not on ART and had a baseline CSF protein <100 mg/dL (150/333) compared to CSF protein ≥100 mg/dL (44/132) (p=0.02), while mortality through 18-weeks was similar in individuals on ART and CSF protein (44/132) (p=0.02) “Fig 1b”.

**Figure 1.**
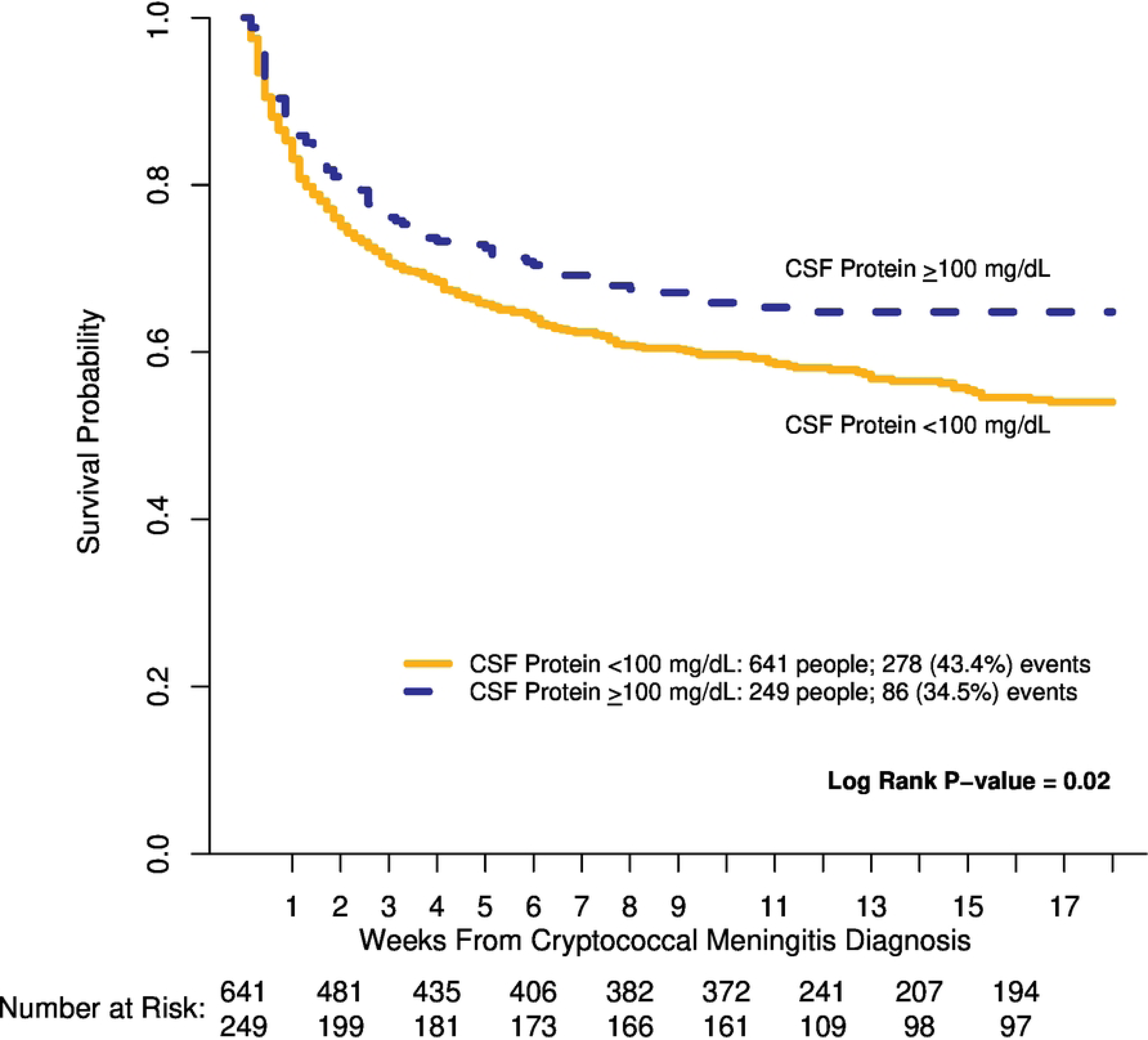
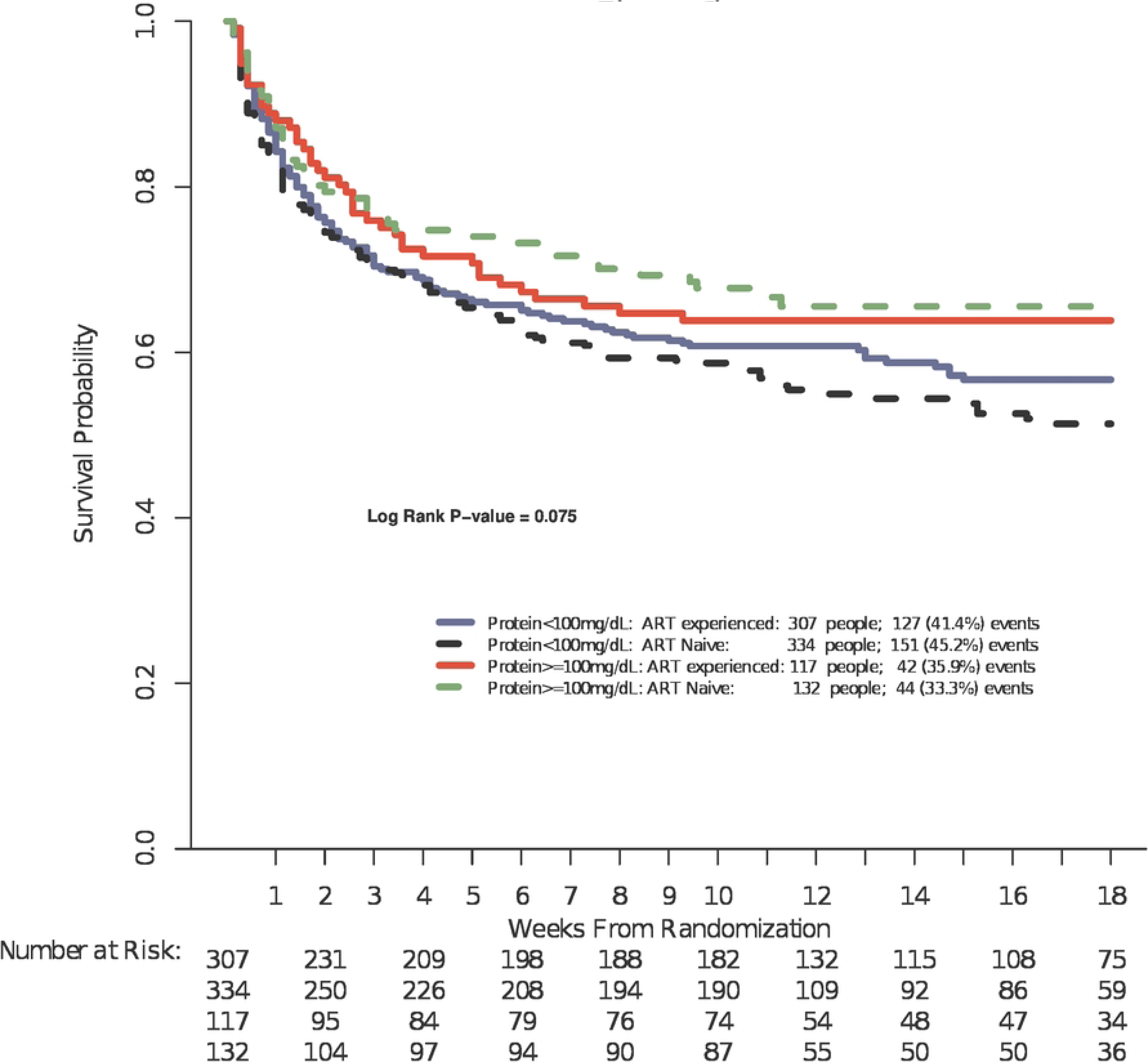
Kaplan-Meier curves of 18-week survival by baseline CSF protein levels alone (1a) and by baseline CSF protein levels and ART status (1b)

**Table 4.**
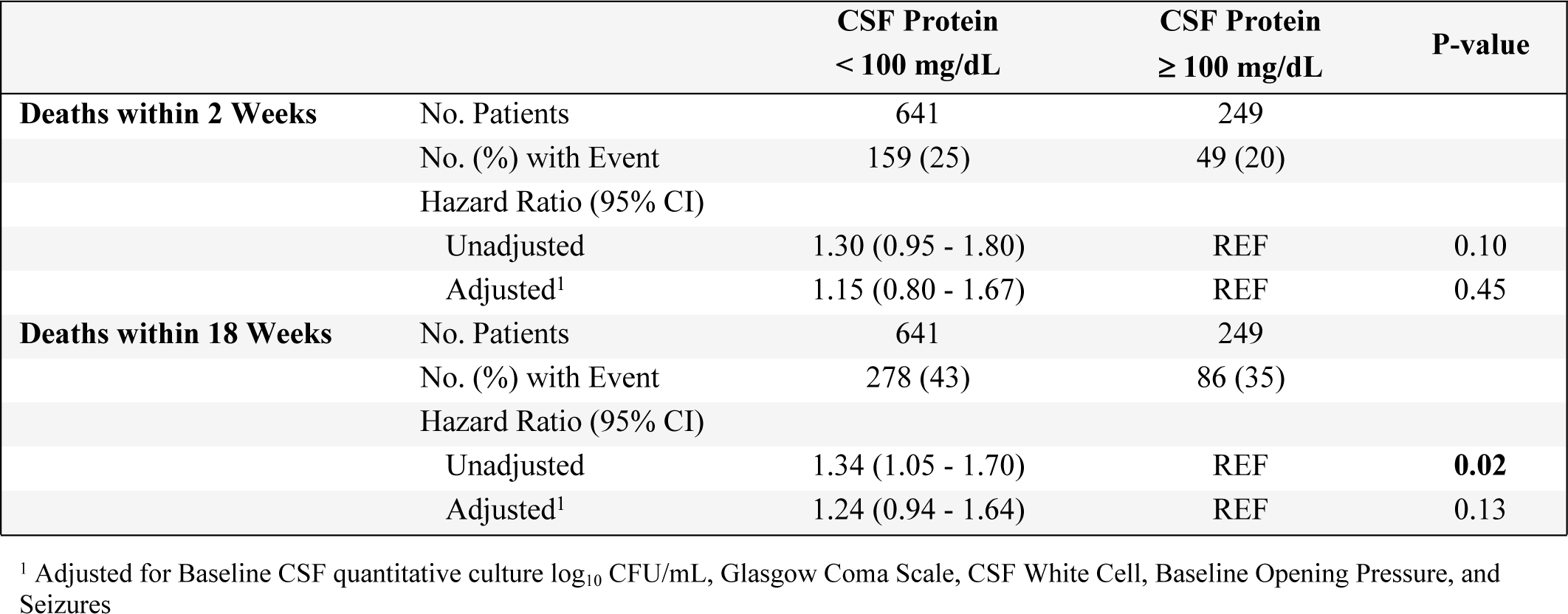
Mortality by Baseline CSF Protein Levels.

|  |  | CSF Protein<br>< 100 mg/dL | CSF Protein<br>≥ 100 mg/dL | P-value |
| --- | --- | --- | --- | --- |
| **Deaths within 2 Weeks** | No. Patients | 641 | 249 |  |
|  | No. (%) with Event | 159 (25) | 49 (20) |  |
|  | Hazard Ratio (95% CI) |  |  |  |
|  | Unadjusted | 1.30 (0.95 - 1.80) | REF | 0.10 |
|  | Adjusted<sup>1</sup> | 1.15 (0.80 - 1.67) | REF | 0.45 |
| **Deaths within 18 Weeks** | No. Patients | 641 | 249 |  |
|  | No. (%) with Event | 278 (43) | 86 (35) |  |
|  | Hazard Ratio (95% CI) |  |  |  |
|  | Unadjusted | 1.34 (1.05 - 1.70) | REF | **0.02** |
|  | Adjusted<sup>1</sup> | 1.24 (0.94 - 1.64) | REF | 0.13 |

<sup>1</sup> Adjusted for Baseline CSF quantitative culture log<sub>10</sub> CFU/mL, Glasgow Coma Scale, CSF White Cell, Baseline Opening Pressure, and Seizures

## Discussion

We demonstrate that in cryptococcal meningitis, persons presenting with CSF protein ≥100 mg/dL had better overall 18-week survival and more frequently presented with altered mental status, seizures, higher CSF white cell count, and a lower CSF quantitative cryptococcal culture. When stratified by ART status, only individuals receiving ART and a CSF protein ≥100 mg/dL presented more frequently with altered mental status. Furthermore, CSF protein ≥100 mg/dL was associated with higher levels of CSF pro-inflammatory, type-1, and type-3 cytokine response. We found a 34% increased hazard of 18-week mortality among individuals with a baseline CSF protein <100 mg/dL as compared to those with CSF protein ≥100 mg/dL. After adjusting for CSF fungal burden, Glasgow coma scale score <15, CSF white cell count, CSF opening pressure, and baseline self-reported seizures, the 18-week mortality difference was only partially attenuated with a remaining 24% increased hazard in mortality that was not statistically different but unexplained by traditional risk factors. Taken together, these findings suggest that CSF protein is a possible predictive biomarker of cryptococcal meningitis severity that is easily accessible at baseline unlike quantitative CSF fungal cultures, which take 7 or more days and are not performed in most routine clinical care settings.

In cryptococcal meningitis, high baseline CSF fungal burden and slow rate of fungal clearance are independent predictors of increased mortality at 2 and 10 weeks [10, 12, 17]. Baseline CSF fungal burden and rate of fungal clearance are influenced by the degree of cell-mediated immunity and host inflammatory response in cryptococcal meningitis [18]. Higher CD4 T-cell count, CSF white cell count, and CSF pro-inflammatory cytokines, IL-6, interferon-γ, and TNF-α, are associated with a lower CSF fungal burden, a more rapid rate of fungal clearance, and improved survival [11, 18-20]. We found that individuals with a baseline CSF protein ≥100 mg/dL presented with a higher CD4 T-cell count and a higher CSF white blood cell count, suggesting the presence of a robust inflammatory response compared to those with a CSF protein <100 mg/dL, even in the absence of ART.

In murine models of C*ryptococcus neoformans* infection, recruitment of CD4^+^ and CD8^+^ leukocytes coincided with CSF cytokine production, white cell recruitment and fungal clearance [21]. In the current study, individuals with CSF protein ≥100 mg/dL exhibited a predominantly pro-inflammatory cytokine response skewed towards type-1 and type-3 T helper cell immune responses. These individuals also exhibited higher levels of some type-2 T helper cell and immune-modulatory cytokines; but this was to a lesser extent (<50% of cytokines in these categories were statistically significant). The elevated levels of chemokines, CXCL1, CXCL2, CXCL8, and CCL20, responsible for T cells, B cells, dendritic cells, monocytes, and neutrophils recruitment, would explain the higher CSF white cell count observed in those with CSF protein ≥100 mg/dL [11, 18, 20, 22-25]. Moreover, we noted increased expression of the immuno-regulatory cytokines, IL-1ra and IL-2, and the immune exhaustion protein PD-L1, which suppress activated T-cells, in those with CSF protein ≥100 mg/dL, suggesting a possible attenuation of damaging inflammatory effects of activated T-cells [21, 26-28]. The combination of this cytokine and cellular response, along with the anti-cryptococcal activity of Granzyme B, likely explains the lower baseline CSF fungal burden and a higher percentage of sterile CSF cultures at day 14 among individuals with CSF protein ≥100 mg/dL [18, 29, 30].

Cell-mediated immunity plays a crucial role in protecting against the development of cryptococcal disease and promoting fungal clearance [21, 31]. Deficiency in CD4 T-cells increases the susceptibility to cryptococcal meningitis and is associated with cryptococcal-related mortality [31, 32]. While most cases of HIV-associated cryptococcal meningitis occur when CD4 T-cell counts drop below 100 cells/μL, approximately 10-20% of cryptococcosis occurs at higher CD4 T-cell counts [32]. Higher CD4 T-cell counts are more frequently associated with altered mental status and lower CSF burden of *Cryptococcus* [32]. Despite low median CD4 T-cell counts in both CSF protein groups, corresponding to advanced HIV disease, individuals with CSF protein ≥100 mg/dL had a statistically significant higher baseline CD4 T-cell count, suggesting the possibility of an enhanced T-cell mediated immune response to *Cryptococcus*. The enhanced T-cell mediated immune response and presence of increased levels of CSF cytokines when CSF protein ≥100 mg/dL may explain the resulting lower baseline CSF fungal burden, higher percentage of sterile cultures at 2 weeks, as well as the occurrence of altered mental status and seizures at baseline. Ultimately, additional research is needed to thoroughly investigate the functionality of CD4 T-cells in the context CSF protein in cryptococcal meningitis.

Despite the higher CSF white cell count, lower CSF fungal burden, and higher levels of CSF cytokines in individuals with a CSF protein ≥100 mg/dL, the survival advantage was negated after adjusting for established risk factors of cryptococcal mortality. Notably, altered mental status was observed in 46% of individuals with a CSF protein ≥100 mg/dL at baseline. We have previously demonstrated that altered mental status in cryptococcosis is most likely associated with a dysfunctional host immune response [33]. Here, we demonstrate a robust inflammatory response in individuals with CSF protein ≥100 mg/dL, characterized by a predominantly pro-inflammatory cytokine response skewed toward type-1 T helper cell and type-3 immune responses. Taken together, our findings suggest that individuals with cryptococcal meningitis presenting with altered mental status and a baseline CSF protein ≥100 mg/dL may be exhibiting a robust inflammatory response leading to neurologic deterioration and death [21]. This is consistent with the damage-response framework proposed by Pirofski and Casadevall, a strong host-mediated inflammatory response during cryptococcal infection can inflict host damage, neurological impairment, and death [34].

Our study has several limitations. Our study population consisted of only people living with HIV and hence may not be generalizable to cases of non-HIV associated cryptococcal meningitis. We measured CSF cytokine levels only at baseline, and inflammation changes over time [35]. We recognize that cytokine levels can fluctuate over time, and we may not have captured the complete dynamics of the immune response in cryptococcal meningitis. We did not assess serum protein levels, which would have allowed for a better evaluation of the CSF to serum protein ratio. The ratio provides information about the permeability and integrity of the blood-brain barrier during meningeal inflammation, which would affect CSF protein level [36]. Lastly, our study combined data from two separate clinical trials which may affect the internal validity of the study.

In conclusion, we have demonstrated that baseline CSF protein levels can serve as a surrogate marker of immune activation in cryptococcal meningitis and provide valuable information on disease severity. Individuals with CSF protein ≥100 mg/dL exhibited a more robust immune response against cryptococcal disease, as indicated by a lower CSF fungal burden at baseline and a higher percentage of sterile CSF cultures at day 14. On the other hand, individuals with a CSF protein <100 mg/dL, particularly those not on ART, demonstrated a deficient or absent cell-mediated immune response, manifested by a higher CSF fungal burden at baseline, lower CSF white blood cell count, fewer sterile cultures at day 14, and higher 18-week morality. Further studies are warranted to investigate the potential of CSF protein levels as a rapid predictive tool for identifying individuals who would benefit from targeted immunomodulatory therapy to either dampen an exaggerated immune response or augment a deficient immune response.

## Data Availability

All relevant data are within the manuscript and its Supporting Information files.

## Acknowledgements

ASTRO-cm and AMBITION-cm study Team Members. Reuben Kiggundu, Andrew Akampurira, Paul Kirumira, Jane Francis Ndyetukira, Cynthia Ahimbisibwe, Florence Kugonza, Carolyne Namuju, Alisat Sadiq, Tadeo Kiiza Kandole, Tony Luggya, Julian Kaboggoza, Eva Laker, Alice Namudde, Sarah Lofgren, Richard Kwizera, Irene Rwomushana, Mike Ssemusu, Joan Rukundo, James Mwesigye, Kirsten Nielsen, Anna Stadelman, Ananta S. Bangdiwala, David Lawrence and Nabila Youssouf.

## Financial support

This work was supported by the National Institute of Neurologic Diseases and Stroke (R01NS086312, K23NS122601); the Fogarty International Center (K01TW010268, R25TW009345); the National Institute of Allergy and Infectious Diseases (T32AI055433); United Kingdom Medical Research Council / DfID / Wellcome Trust Global Clinical Trials (grant numbers M007413/1, MR/P006922/1); Grand Challenges Canada (S4-0296-01); and the European and Developing Countries Clinical Trials Partnership (EDCTP) (TRIA2015-1092) with assistance from the Swedish International Development Cooperation Agency (SIDA), the U.K. Department of Health and Social Care, the U.K. Foreign Commonwealth and Development Office, and the U.K. Medical Research Council.

## Potential conflicts of interest

All authors: no reported conflicts of interest.

## Notes

### Competing Interest Statement

The authors have declared that no competing interests exist.

### Funding Statement

The author(s) received no specific funding for this work.

